# Pre-existing Cardiovascular Disease in United States Population at High Risk for Severe COVID-19 Infection

**DOI:** 10.1101/2020.05.11.20089714

**Authors:** Adnan I. Qureshi, Wei Huang, Iryna Lobanova, Farhan Siddiq, M. Fareed K. Suri, S. Hasan Naqvi, Brandi R. French, Camilo R. Gomez

## Abstract

**Background and Purpose:** There is increasing recognition of a relatively high burden of pre-existing cardiovascular disease in Corona Virus Disease 2019 (COVID-19) infected patients. We determined the burden of pre-existing cardiovascular disease in persons residing in United States (US) who are at risk for severe COVID-19 infection.

**Methods:** Age (≥60 years), presence of chronic obstructive pulmonary disease, diabetes, mellitus, hypertension, and/or malignancy were used to identify persons at risk for admission to intensive care unit, or invasive ventilation, or death with COVID-19 infection. Persons were classified as low risk (no risk factors), moderate risk (1 risk factor), and high risk (two or more risk factors present) using nationally representative sample of US adults from National Health and Nutrition Examination Survey 2017-2018 survey.

**Results:** Among a total of 5856 participants, 2386 (40.7%) were considered low risk, 1325 (22.6%) moderate risk, and 2145 persons (36.6%) as high risk for severe COVID-19 infection. The proportion of patients who had pre-existing stroke increased from 0.6% to 10.5% in low risk patients to high risk patients (odds ratio [OR]19.9, 95% confidence interval [CI]11.6-34.3). The proportion of who had pre-existing myocardial infection (MI) increased from 0.4% to 10.4% in low risk patients to high risk patients (OR 30.6, 95% CI 15.7-59.8).

**Conclusions:** A large proportion of persons in US who are at risk for developing severe COVID-19 infection are expected to have pre-existing cardiovascular disease. Further studies need to identify whether targeted strategies towards cardiovascular diseases can reduce the mortality in COVID-19 infected patients.

---

Pre-existing cardiovascular disease has been identified in 8-15% of patients with Corona Virus Disease 2019 (COVID-19) infection who were hospitalized in China^1, 2^ and increased ICU admission and in hospital mortality. ^2^ The existing data from China but may not be applicable to United States (US) due to lower proportion of hypertension and dyslipidemia and higher proportion of cigarette smoking in Chinese population.^3^ Knowledge of burden of pre-existing cardiovascular disease in persons at risk for severe COVID-19 infection may be important to determine the cardiovascular needs due to rapid increase in COVID-19 infected patients in US. We performed this analysis to examine the prevalence of pre-existing cardiovascular disease in patients at high risk of severe COVID-19 infection defined by admission to intensive care unit, or invasive ventilation, or death in a nationally representative sample of the US population.

## METHODS

### Defining persons at high risk for severe COVID-19 infection

We dichotomized age as <60 and ≥60 years for identifying patients at risk for severe COVID-19 infection (ICU admission and mortality) based on a previous study.^2^ Guan et al.^4^ identified presence of chronic obstructive pulmonary disease, diabetes, mellitus, hypertension, or malignancy as predictors for composite endpoint of admission to intensive care unit, or invasive ventilation, or death among 1590 COVID-19 infected patients who were hospitalized in one of the 575 hospitals within China. The risk was higher if 2 or more risk factors were present in the same individual.

### US nationally representative data

We used the most recent nationally representative sample of US adults from population screened in the National Health and Nutrition Examination Survey (NHANES) 2017-2018 survey cycle. ^5^ The NHANES represents the noninstitutionalized civilian resident population of US. Persons are derived from a stratified multistage probability sample. Data was collected by directly interviewing survey participants and by conducting clinical tests and physical examinations.

### Identification of high risk for severe COVID-19 infection

We included persons aged ≥18 years in the analysis. Presence of chronic obstructive pulmonary disease, diabetes mellitus, hypertension, and malignancy was ascertained by reviewing answers to query during interview if a doctor or other health professional had told them if they had the abovementioned diseases. Presence of hypertension was also identified if the mean of 4 properly measured BP readings was equal or greater than 140/90 mm Hg. Diabetes mellitus was also identified if HbA1c, glycated hemoglobin was greater than 6.4%. Persons were classified as low risk (no risk factors), moderate risk (1 risk factor), and high risk (two or more risk factors present).

### Identification of pre-existing cardiovascular disease

Persons were considered as having had a stroke or myocardial infarction (MI) if they reported that they had been told by a physician that they had suffered a stroke or heart attack.

### Statistical analysis

The analysis was descriptive and proportion (percentage with or without 95% confidence interval [CI]) of persons with pre-existing stroke and MI were presented. The crude odd ratio (OR) with 95% CI was calculated when relevant.

## RESULTS

Among a total of 5856 participants, 2386 (40.7%) were considered low risk, 1325 (22.6%) moderate risk, and 2145 persons (36.6%) as high risk for severe COVID-19 infection. The proportion of patients who had pre-existing stroke increased from 0.6% to 10.5% in low risk patients to high risk patients (OR 19.9, 95% CI 11.6-34.3, Table 1). The proportion of who had pre-existing MI increased from 0.4% to 10.4% in low risk patients to high risk patients (OR 30.6, 95% CI 15.7-59.8). The distribution of persons with pre-existing stroke or MI in various strata (gender and race/ethnicity) is presented in Tables 2 and 3, respectively. The highest burden of preexisting stroke was seen in non-Hispanic black persons and non-Hispanic white persons with lowest prevalence seen in Mexican Americans and non-Hispanic Asian in high risk strata. The highest burden of preexisting MI was seen in non-Hispanic white persons. There was a higher proportion of men (compared with women) with preexisting MI in high risk strata.

**Table 1.** The proportion of US persons with pre-existing stroke and MI according to risk for severe COVID-19 infection: NHANES 2017-2018.

| Risk level for severe COVID-19 infection | Total | Pre-existing stroke |  | Pre-existing MI |  |
| --- | --- | --- | --- | --- | --- |
|  |  | Proportion with stroke (95% CI) | Odds ratio (95% CI) | Proportion with MI (95 % CI) | Odds ratio (95% CI) |
| Low risk | 2386(40.7%) | 0.6% (0.3%-0.9%) | 1(1-1) | 0.4% (0.1%-0.6%) | 1(1-1) |
| Moderate risk | 1325(22.6%) | 2.5% (1.7%-3.3%) | 4.3(2.3-8.1) | 2.9% (2.0%-3.8%) | 7.8 (3.8-16.1) |
| High risk | 2145(36.6%) | 10.5% (9.2%-11.8%) | 19.9(11.6-34.3) | 10.4% (9.1%-11.7%) | 30.6 (15.7-59.8) |

**Table 2.** The proportion of US persons with pre-existing stroke in various gender and race/ethnicity strata according to risk for severe COVID-19 infection-NHANES 2017-2018

| Risk level for severe COVID-19 infection | Gender |  | Race/ethnicity |  |  |  |  |  |
| --- | --- | --- | --- | --- | --- | --- | --- | --- |
|  | Men | Women | Mexican American | Other Hispanic | Non-Hispanic White | Non-Hispanic Black | Non-Hispanic Asian | Other Race |
| Low risk | 0.6% | 0.5% | 0.2% | 0% | 0.7% | 1.1% | 0.2% | 1.6% |
| Moderate risk | 2.2% | 2.8% | 2.8% | 3.4% | 2.2% | 3.9% | 0.5% | 1.3% |
| High risk | 10.7% | 10.4% | 4.8% | 4.9% | 12.1% | 12.9% | 4.3% | 20.2% |

**Table 3.** The proportion of US persons with pre-existing MI in various gender and race/ethnicity strata according to risk for severe COVID-19 infection-NHANES 2017-2018

| Risk level for severe COVID-19 infection | Gender |  | Race/ethnicity |  |  |  |  |  |
| --- | --- | --- | --- | --- | --- | --- | --- | --- |
|  | Men | Women | Mexican American | Other Hispanic | Non-Hispanic White | Non-Hispanic Black | Non-Hispanic Asian | Other Race |
| Low risk | 0.6% | 0.2% | 0.2% | 0% | 0.7% | 0.2% | 0.2% | 0.8% |
| Moderate risk | 3.8% | 1.9% | 2.8% | 3.4% | 3.3% | 3.3% | 0.5% | 3.8% |
| High risk | 14.8% | 6.0% | 7.6% | 8.8% | 14.0% | 7.1% | 6.1% | 17.0% |

## DISCUSSION

The relatively high prevalence (approximately 10%) of either pre-existing stroke or MI in persons in US at risk for severe COVID-19 infection needs to be recognized. The odds of either pre-existing stroke or MI was 19 to 30 folds higher in high risk persons compared with low risk persons. The United States demographic data suggest that there are 209,128,094 adults (aged ≥18 years) residing in United States^6^. Assuming 36.6% (76,540,882 persons) of the US adult population is high risk, and 10.5% of those who would either have pre-existing stroke or MI, 8,036,793 persons in US may acquire severe COVID-19 infection with pre-existing cardiovascular disease. The reason for concurrent existence is probably related to common risk factors such as old age, hypertension, diabetes mellitus, and malignancy.^7^ The burden of pre-existing MI was higher in high risk men than women but no difference was noted in burden of pre-existing stroke in high risk men than women. The burden was disproportionate between various race/ethnic groups with highest prevalence in non-Hispanic white persons.

The definitions of MI and stroke were based on self-reported physician diagnoses. Self-report for history of stroke has a sensitivity and specificity of 95% and 96%, respectively.^8^ Self-report of prevalent MI has a high sensitivity (74% to 100%) and specificity (94% to 99%).^9, 10^ Previous studies have suggested that self-reported diagnoses in the NHANES surveys is reasonably accurate.^11^ A study^12^ found interview reports using the same question for MI as that used in NHANES 2017-2018 had a true positive rate of 83% when compared with hospitalization records in 10,523 participants from NHANES I. The study also noted high specificity observed for self-reported hypertension, diabetes mellitus, and hyperlipidemia.

A large proportion of persons in US who are at risk for developing severe COVID-19 infection are expected to have pre-existing cardiovascular disease. Pre-existing cardiovascular disease increased the risk higher risk of myocardial injury^13^ and new stroke^14^ and in hospital mortality^13, 14^ in COVID-19 infected patients. Further studies need to identify whether targeted strategies towards cardiovascular diseases in such patients can reduce the mortality in COVID-19 infected patients.

## Data Availability

The data is available through NHANES website

## References

1. Shi, H, Han, X, Jiang, N, Cao, Y, Alwalid, O, Gu, J, et al., Radiological findings from 81 patients with COVID-19 pneumonia in Wuhan, China: a descriptive study. Lancet Infect Dis, 2020. 20(4): p. 425–434.

2. Zhou, F, Yu, T, Du, R, Fan, G, Liu, Y, Liu, Z, et al., Clinical course and risk factors for mortality of adult inpatients with COVID-19 in Wuhan, China: a retrospective cohort study. Lancet, 2020. 395(10229): p. 1054-1062.

3. Kuulasmaa, K, Tunstall-Pedoe, H, Dobson, A, Fortmann, S, Sans, S, Tolonen, H, et al., Estimation of contribution of changes in classic risk factors to trends in coronary-event rates across the WHO MONICA Project populations. Lancet, 2000. 355(9205): p. 675–87.

4. Guan WJ, Liang WH, Zhao Y, et al. Comorbidity and its impact on 1590 patients with Covid-19 in China: A Nationwide Analysis [published online ahead of print, 2020 Mar 26]. EurRespirJ. 2020;2000547. doi:10.1183/13993003.00547-2020

5. NHANES 2017–2018 Overview. 2020 February 21, 2020; Available from: https://www.n.cdc.gov/nchs/nhanes/continuousnhanes/overview.aspx?BeginYear=2017.

6. United States Demographic Statistics. Available from: https://www.infoplease.com/us/comprehensive-census-data-state/demographic-statistics-342.

7. Khot, UN, Khot, MB, Bajzer, CT, Sapp, SK, Ohman, EM, Brener, SJ, et al., Prevalence of conventional risk factors in patients with coronary heart disease. JAMA, 2003. 290(7): p. 898–904.

8. O’Mahony, PG, Dobson, R, Rodgers, H, James, OF, and Thomson, RG, Validation of a population screening questionnaire to assess prevalence of stroke. Stroke, 1995. 26(8): p. 1334-7.

9. Hagman, M, Jonsson, D, and Wilhelmsen, L, Prevalence of angina pectoris and myocardial infarction in a general population sample of Swedish men. Acta Med Scand, 1977. 201(6): p. 571–7.

10. Olsson, L, Svardsudd, K, Nilsson, G, Ringqvist, I, and Tibblin, G, Validity of a postal questionnaire with regard to the prevalence of myocardial infarction in a general population sample. Eur Heart J, 1989. 10(11): p. 1011-6.

11. Madow, WG, Net differences in interview data on chronic conditions and information derived from medical records. Vital Health Stat 2, 1973(57): p. 1–58.

12. Bergmann, MM, Byers, T, Freedman, DS, and Mokdad, A, Validity of self-reported diagnoses leading to hospitalization: a comparison of self-reports with hospital records in a prospective study of American adults. Am J Epidemiol, 1998. 147(10): p. 969–77.

13. Guo, T, Fan, Y, Chen, M, Wu, X, Zhang, L, He, T, et al., Cardiovascular Implications of Fatal Outcomes of Patients With Coronavirus Disease 2019 (COVID-19). JAMA Cardiology, 2020. Published online March 27, 2020. doi:10.1001/jamacardio.2020.1105

14. Li, Y, Wang, M, Zhou, Y, Chang, J, Xian, Y, Mao, L, et al., Acute cerebrovascular disease following COVID-19: A Single Center, Retrospective, Observational Study. Available at SSRN: https://ssrn.com/abstract=3550025 or http://dx.doi.org/10.2139/ssm.3550025 [Epub ahead of print]

